# Medical Service Use for Dental Problems in Australia: A National Cross-Sectional Study

**DOI:** 10.64898/2026.09.17.26363370

**Authors:** Liana Luzzi, Kym M. McCormick, Najith Amarasena, Gloria Mejia, Sergio Chrisopoulos

## Abstract

**Aim:** To examine how and why Australians seek medical rather than dental care for dental problems, and to describe the pathways and treatments provided within medical settings.

**Subjects and Methods:** Data were drawn from the National Dental Telephone Interview Survey (NDTIS) 2021, a nationally representative survey of 5,526 adults. Survey-weighted analyses estimated the prevalence and sociodemographic predictors of medical service use for dental problems. Among respondents who sought medical care, we identified the highest-intensity pathway used (general practice/other primary care, emergency department, hospital admission) and described treatment received—including prescription medications, non-pharmaceutical interventions, and referrals—stratified by reason for presentation.

**Results:** Overall, 4.4% of adults sought medical care for a dental problem in the preceding year. Prevalence was substantially higher among Indigenous adults (13.5%), low-income groups (6.5%), and those eligible for public dental care (8.5%). Emergency departments were the dominant pathway (76.9%), followed by hospital admission (13.6%) and primary care (9.5%). Toothache/wisdom-tooth problems and oral injuries had the highest rates of antibiotic prescribing, while procedural care predominated for oral injuries. Other acute dental problems were managed heterogeneously, with many receiving only advice or symptomatic treatment. Referral to a dentist was uncommon across all groups.

**Conclusion:** A significant proportion of Australians rely on medical services for dental problems, particularly those facing financial and geographic barriers to dental care. These encounters frequently provide symptomatic rather than definitive treatment and rarely include referral for follow-up dental care. The findings highlight gaps in access to timely dental services and the need for strengthened integration between medical and dental systems, improved referral mechanisms, and policy interventions to reduce preventable and inappropriate medical presentations for dental conditions.

## INTRODUCTION

Despite substantial improvements in oral health over recent decades, driven by water fluoridation, widespread use of fluoridated toothpaste, and better hygiene practices, dental problems remain prevalent in Australia.

National data indicate that approximately 11% of dentate adults have an inadequate dentition (<21 teeth), more than 95% of adults born before 1970 have experienced dental caries, and one in three adults has untreated decay (Brennan et al., 2020). Gum disease is common, with one in three dentate adults experiencing moderate or severe periodontitis, and dental symptoms remain widespread: in the preceding year, 20% experienced toothache, 27% felt they needed an extraction or filling, and 24% rated their oral health as fair or poor.

Despite this burden, around two thirds of Australians are ineligible for publicly funded dental care and must rely on private dental services (Duckett et al., 2019). Because dental care sits largely outside Medicare, those without private insurance face substantial out-of-pocket costs, while public dental waiting lists remain long and capacity is limited, with geographic maldistribution of the dental workforce further exacerbating barriers to timely care, particularly in rural and remote areas (AIHW, 2023b; Hopcraft, 2024). These structural barriers contribute to delayed dental treatment and increasing clinical severity at the time of presentation.

When individuals cannot access timely or affordable dental care, many seek help from general practitioners or hospital emergency departments. While these providers can offer symptomatic relief (typically pain medication or antibiotics) they are rarely able to provide definitive treatment, often resulting in repeated presentations and escalating disease (Davis et al., 2010). Nationally, an estimated 750,000 medical practitioner encounters occur annually for dental problems, yet the total system cost remains unknown; available estimates vary widely (between $30 and $300 million), reflecting the lack of coordinated national monitoring (National Advisory Council on Dental Health, 2012). In 2020–21, 82,916 potentially preventable hospitalisations for dental conditions were recorded, up from 60,251 in 2009–10, with rates rising from 3.0 per 1,000 population in major cities to 4.6 per 1,000 in very remote areas (AIHW, 2023a). The most recent national cost estimate (now more than a decade old) placed the annual cost of dental hospitalisations at $84 million in 2009–10 (National Advisory Council on Dental Health, 2012), and the financial burden today is almost certainly far greater.

These patterns reflect a system in which failures of access, driven by limited public services, high private costs and geographic inaccessibility, shift dental demand onto medical settings not equipped to provide definitive care (Duckett et al., 2019). Prior research has noted this shift (Hopcraft, 2024; Lee et al., 2012), but nationally representative data remain limited regarding who is affected, what treatments are provided, and whether dental follow-up occurs.

To address this gap, we used nationally representative data to quantify how often Australians seek medical rather than dental care for dental problems and to identify the sociodemographic factors associated with this behaviour. We examined the medical pathways used (including general practice, emergency departments, and hospital admission) and how these varied by presentation type. Among those who sought medical care, we also described the treatments provided, including prescription and non-pharmaceutical management and referral patterns. These analyses provide the first contemporary national picture of how non-dental health services in Australia respond to dental problems, and who is most affected by this pattern of care.

## METHODS

### Study Design and Sample Selection

Data were drawn from the 2021 National Dental Telephone Interview Survey (NDTIS), a cross–sectional survey of adults aged 18 years and over across Australia. A stratified sampling design was used, with selection conducted by Services Australia using the Medicare database as the sampling frame. Strata were defined using the Australian Statistical Geography Standard Greater Capital City Statistical Areas (GCCSA), grouping areas into Greater City or Rest of State within each jurisdiction. Individuals listed on the Medicare database were randomly selected, and data were weighted following standard procedures for stratified random samples. Ethics approval was obtained from the University of Adelaide Human Research Ethics Committee (H–2020–153).

Participants were invited to complete a structured questionnaire via computer–assisted telephone interview (CATI) or online survey.

### Outcome measures

#### Medical service use for a dental problem

The primary outcome was use of medical services for a dental problem, measured by the item: “In the last year, did you visit a health provider, other than a dental practitioner, for a dental problem?” Participants could select one or more service types: hospital emergency department, hospital admission, general practitioner, or other medical provider. Any affirmative selection was coded as 1 = Yes and 0 = No, with cases missing all four items coded as missing.

#### Highest-intensity care pathway

Among participants who used medical services for a dental problem, we also derived a three-level ordered outcome reflecting the highest-intensity service pathway: Primary/Other care, Emergency Department, and Hospital admission, prioritised in that order of intensity.

#### Reason for presentation

Respondents who used a medical service were asked to report their reason for presenting. Responses were grouped into three clinically meaningful categories: Toothache or wisdom-tooth problem; Oral injury; and Other acute dental problem.

#### Treatment and medication received

Participants were asked whether they received a prescription (and whether it was filled) and whether they received any non-pharmaceutical treatment during their visit. Open-text responses describing the *type* of medication or treatment were coded into predefined categories. Medication-related responses were coded as: Pain relief; Antibiotics; Both pain relief and antibiotics; Don’t know / can’t recall. Non-pharmaceutical treatment responses were coded into: Dental-type treatment (e.g., drainage, irrigation, dressing, stabilisation); Other medical treatment (e.g., wound care, supportive management); Referral to a dentist; Other treatment; and No treatment. Coding followed a structured deductive framework. Ambiguous responses were independently reviewed by two authors (SC and KM), with discrepancies resolved through consensus.

### Explanatory Variables

Seven explanatory variables were selected a priori based on their relevance to access-to-care research: age, sex, Indigenous identity, educational attainment, household income, service coverage, and residential location. Age was coded into five ordered groups (18–24, 25–44, 45–59, 60–74, and 75+ years). Indigenous identity was assessed using the question: *“Do you identify as being of Aboriginal or Torres Strait Islander origin?”* Responses indicating Aboriginal and/or Torres Strait Islander identity were combined into a single Indigenous category for analysis. Educational attainment was derived from two survey items on highest school year completed and highest qualification, classified through a hierarchical rule-set into: university degree or higher; certificate/diploma; Year 12; Year 10–11; Year 9 or below; and currently studying (no qualification). Household income was grouped into four ordered categories: <$50,000; $50,000–<$100,000; $100,000–<$150,000; and ≥$150,000. Service coverage was derived by combining private dental insurance status with eligibility for public dental care, producing four categories that reflect real-world access contexts: (1) private insurance only, (2) eligible for public dental care only, (3) both private insurance and public eligibility, and (4) no insurance / not eligible. Two alternative coding schemes (conservative vs. liberal) were evaluated; the conservative scheme produced more stable classification and was retained for analysis. Finally, residential location was coded into major cities, regional (inner and outer regional), and remote/very remote areas.

Non-substantive responses (“don’t know/rather not say”, “not applicable”, “not stated”) were retained in descriptive analyses but excluded from regression models.

### Missingness and imputation

Missingness was primarily due to explicit non-substantive income responses (~15%). Smaller amounts of missingness occurred for insurance access group (4%) and education attainment (0.8%); all remaining variables were fully observed. To minimise bias and maximise statistical efficiency, missing data were handled through multiple imputation by chained equations (Rubin, 1996; Sterne et al., 2009) implemented in R using the mice package (Van Buuren & Groothuis-Oudshoorn, 2011). One hundred imputed datasets were generated with 20 iterations per chain. Diagnostic checks indicated satisfactory convergence: trace plots demonstrated stable, well-mixed chains; Monte Carlo error was <10% of the corresponding standard errors; and the fraction of missing information for income variables ranged from 0.09–0.13. Strip plots confirmed the plausibility of imputed values relative to observed distributions. Imputed datasets were used for all regression analyses.

### Data Analysis

#### Descriptive analyses

Survey-weighted prevalence estimates and 95% confidence intervals (CIs) were obtained using Taylor-linearised variance estimation, with replicate-weight bootstrap standard errors (Lumley, 2004) being applied where sparse cells increased instability. Following standard disclosure rules, cells with fewer than five unweighted observations were suppressed. Care pathways (GP/Other, ED, Admission) were summarised descriptively across demographic, socioeconomic, and access-to-care variables. Treatment patterns were examined overall and stratified by reason for presentation (toothache/wisdom tooth, oral injury, other acute problem).

#### Regression analyses

Associations between each explanatory variable and the likelihood of using medical services for a dental problem were examined using separate survey-weighted quasi-Poisson regression models with a log link. Each model adjusted for age and sex only (i.e., *Outcome ~ age + sex + explanatory variable*). This approach directly estimates relative risks while accommodating mild overdispersion and remaining compatible with design-based variance estimation (Lumley, 2004; Zou, 2004). Regression modelling was not carried out for care pathways or treatment categories due to small subgroup sample sizes and the descriptive purpose of those analyses. All analyses were conducted in R version 4.4.2 using the *survey* and *mice* packages.

## RESULTS

### Sample Characteristics and Overall Prevalence

A total of 5,526 adults participated in the 2021 NDTIS (response rate: 24.8%). After weighting, the sample was 56.4% female with a mean age of 48.9 years (SD = 17.6). Overall, 4.4% of adults reported using a medical service for a dental problem in the previous 12 months (Table 1). Although uncommon in the general population, prevalence differed substantially across sociodemographic groups.

**Table 1.** Prevalence and Predictors of Medical Service Use for Dental Problems: Survey-Weighted Estimates and Poisson Regression.

| Characteristic | n | % (95% CI) | aPR (95% CI) |
| --- | --- | --- | --- |
| **All** | **5,333** | **4.4% (3.5%–5.2%)** | **-** |
| **Sex** |  |  |  |
| Male | 2,325 | 3.9% (2.6%–5.1%) | Ref |
| Female | 3,008 | 4.8% (3.7%–6.0%) | 1.25 (0.85, 1.85) |
| **Age** |  |  |  |
| 18–24 | 448 | 5.5% (2.7%–8.3%) | 0.67 (0.37, 1.20) |
| 25–44 | 1,938 | 3.7% (2.6%–4.8%) | 0.78 (0.41, 1.48) |
| 45–59 | 1,273 | 4.3% (2.6%–6.1%) | Ref |
| 60–74 | 1,265 | 5.8% (3.1%–8.5%) | 1.06 (0.53, 2.11) |
| 75+ | 409 | 2.4% (0.2%–4.6%) | 0.45 (0.16, 1.28) |
| **Indigenous Identity** |  |  |  |
| Non-Indigenous | 5,196 | 4.1% (3.3%–5.0%) | Ref |
| Indigenous * | 137 | **13.5% (5.8%–21.2%)** | **3.66 (2.01–6.66)** |
| **Household Income** |  |  |  |
| Less than \$50,000 | 1,179 | **6.5% (4.4%–8.6%)** | **2.72 (1.29–5.72)** |
| \$50,000 to < \$100,000 | 1,276 | 4.5% (2.8%–6.2%) | 1.88 (0.91–3.90) |
| \$100,000 to < \$150,000 | 878 | 3.2% (1.5%–4.8%) | 1.34 (0.57–3.15) |
| Greater than \$150,000 | 1,191 | 2.3% (1.0%–3.7%) | Ref |
| *Don't know / rather not say* † | 775 | 5.1% (2.3%–7.8%) | - |
| *Not stated* † | 34 | 10.8% (0.0%–27.7%) | - |
| **Highest Education Attainment** |  |  |  |
| University degree or higher | 1,110 | 3.4% (2.0%–4.9%) | Ref |
| Certificate/Diploma (non-degree) | 2,969 | 4.9% (3.5%–6.2%) | 1.42 (0.85–2.37) |
| Year 12 or less | 1,005 | 3.4% (1.8%–5.0%) | 0.94 (0.49–1.78) |
| Currently studying (no qualification) * | 207 | 8.2% (3.4%–13.1%) | 2.79 (0.96–8.16) |
| *Don't know / rather not say / not stated* † | 42 | 2.6% (0.0%–7.8%) | - |
| **Service coverage** |  |  |  |
| Private dental insurance only | 2,503 | 3.5% (2.1%–4.8%) | Ref |
| Both private insurance and public eligibility | 485 | 2.7% (1.0%–4.5%) | 0.89 (0.39, 2.07) |
| Eligible for public dental care only | 758 | **8.5% (5.8%–11.2%)** | **2.66 (1.50, 4.71)** |
| No insurance / not eligible | 1,376 | 4.3% (2.8%–5.9%) | 1.25 (0.75, 2.08) |
| *Don't know / rather not say / not stated* † | 211 | 3.7% (0.0%–8.2%) | - |
| **Residential Location** |  |  |  |
| Major cities | 3,163 | 4.2% (3.1%–5.3%) | Ref |
| Regional | 1,946 | 4.7% (3.4%–6.0%) | 1.12 (0.76–1.65) |
| Remote/Very remote * | 224 | 7.5% (1.0%–13.9%) | 1.80 (0.73–4.42) |

**Note.** Sample size (n) reflects the unweighted number of respondents. Percentages represent population-weighted estimates with 95% confidence intervals derived from Taylor-linearised standard errors, accounting for the complex survey design. Estimates were obtained from multiply imputed datasets using survey-weighted quasi-Poisson regression with a log link and Taylor-linearised (robust) standard errors; exponentiated coefficients represent adjusted prevalence ratios (aPRs). Each model included age group and sex as covariates. Values above 1 indicate higher relative risk compared with the reference category (Ref); values below 1 indicate lower risk. Boldface indicates aPRs where the 95% CI does not include 1.00.
\* Precision of estimates varied across subgroups; wide confidence intervals, particularly for groups with smaller unweighted sample sizes, indicate greater uncertainty. Interpretations should focus on both point estimates and CIs.
† “Don’t know / rather not say / not stated” are shown in the descriptive estimates but were excluded from regression analyses; aPRs are therefore reported only for substantive response categories.

Females reported slightly higher use than males (4.8% vs. 3.9%), although the age- and sex-adjusted estimate (aPR = 1.25; 95% CI: 0.85–1.85) indicated considerable uncertainty around this difference. Prevalence varied by age but estimates were imprecise, with the highest levels observed among adults aged 60–74 years (5.8%) and 18–24 years (5.5%), and the lowest among adults aged 75 years and older (2.4%).

### Sociodemographic Associations of Medical Service Use

Marked disparities were observed across socioeconomic and population groups. Indigenous adults had more than three times the adjusted rate of medical service use compared with non-Indigenous adults (13.5% vs. 4.1%; aPR = 3.66; 95% CI: 2.01–6.66). A clear income gradient was evident: adults in the lowest household-income category (<$50,000) had the highest prevalence (6.5%) and nearly triple the adjusted prevalence of those earning ≥$150,000 (aPR = 2.72; 95% CI: 1.29–5.72). Service coverage showed strong associations. Adults eligible for public dental care only had the highest prevalence (8.5%; aPR = 2.66; 95% CI: 1.50–4.71), whereas those with both private insurance and public eligibility had the lowest (2.7%). Educational attainment showed no consistent pattern, and differences across residential location were modest and imprecise; although prevalence reached 7.5% in remote/very remote areas, confidence intervals were wide.

### Medical Care Pathways

Among adults who sought medical care for a dental problem (n = 210), most were managed in emergency departments (76.9%) as their highest intensity point of care. Only 9.5% were managed solely in primary/other care, and 13.6% required hospital admission (Table 2). Pathways differed across population groups. Females had a higher proportion of hospital admissions (17.9%) than males (7.9%). Younger adults (18–24 years) were more likely to be managed in primary care (23.3%), whereas adults aged 60–74 years almost exclusively used ED services (91.4%). Substantial inequities were observed for Indigenous adults, one quarter of whom had a hospital admission (25.7%) compared with 12.5% of non-Indigenous adults. Income was also associated with escalation of care: adults in the lowest income group had the highest admission rates (20.1%). Those eligible for public dental care only also experienced higher admission rates (22.8%). Geographic gradients were pronounced, with hospital admissions accounting for 61.7% of medical use presentations in remote/very remote areas, though estimates were imprecise due to small cell sizes.

**Table 2.**
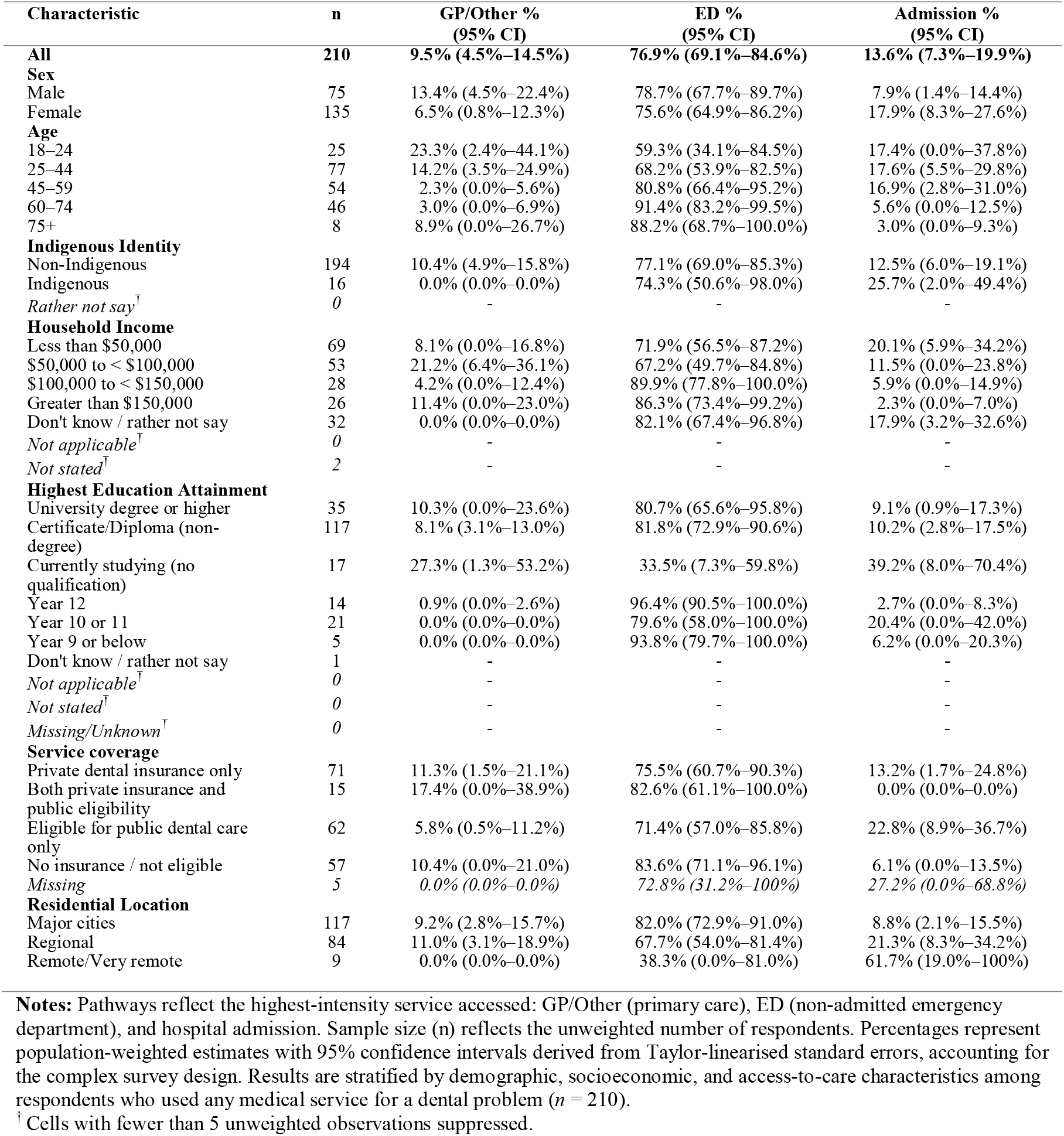
Care pathways for medical presentations for dental problems: survey-weighted proportions and 95% confidence intervals.

| Characteristic | n | GP/Other %<br>(95% CI) | ED %<br>(95% CI) | Admission %<br>(95% CI) |
| --- | --- | --- | --- | --- |
| **All** | **210** | **9.5% (4.5%–14.5%)** | **76.9% (69.1%–84.6%)** | **13.6% (7.3%–19.9%)** |
| **Sex** |  |  |  |  |
| Male | 75 | 13.4% (4.5%–22.4%) | 78.7% (67.7%–89.7%) | 7.9% (1.4%–14.4%) |
| Female | 135 | 6.5% (0.8%–12.3%) | 75.6% (64.9%–86.2%) | 17.9% (8.3%–27.6%) |
| **Age** |  |  |  |  |
| 18–24 | 25 | 23.3% (2.4%–44.1%) | 59.3% (34.1%–84.5%) | 17.4% (0.0%–37.8%) |
| 25–44 | 77 | 14.2% (3.5%–24.9%) | 68.2% (53.9%–82.5%) | 17.6% (5.5%–29.8%) |
| 45–59 | 54 | 2.3% (0.0%–5.6%) | 80.8% (66.4%–95.2%) | 16.9% (2.8%–31.0%) |
| 60–74 | 46 | 3.0% (0.0%–6.9%) | 91.4% (83.2%–99.5%) | 5.6% (0.0%–12.5%) |
| 75+ | 8 | 8.9% (0.0%–26.7%) | 88.2% (68.7%–100.0%) | 3.0% (0.0%–9.3%) |
| **Indigenous Identity** |  |  |  |  |
| Non-Indigenous | 194 | 10.4% (4.9%–15.8%) | 77.1% (69.0%–85.3%) | 12.5% (6.0%–19.1%) |
| Indigenous | 16 | 0.0% (0.0%–0.0%) | 74.3% (50.6%–98.0%) | 25.7% (2.0%–49.4%) |
| Rather not say<sup>†</sup> | 0 | - | - | - |
| **Household Income** |  |  |  |  |
| Less than \$50,000 | 69 | 8.1% (0.0%–16.8%) | 71.9% (56.5%–87.2%) | 20.1% (5.9%–34.2%) |
| \$50,000 to < \$100,000 | 53 | 21.2% (6.4%–36.1%) | 67.2% (49.7%–84.8%) | 11.5% (0.0%–23.8%) |
| \$100,000 to < \$150,000 | 28 | 4.2% (0.0%–12.4%) | 89.9% (77.8%–100.0%) | 5.9% (0.0%–14.9%) |
| Greater than \$150,000 | 26 | 11.4% (0.0%–23.0%) | 86.3% (73.4%–99.2%) | 2.3% (0.0%–7.0%) |
| Don't know / rather not say | 32 | 0.0% (0.0%–0.0%) | 82.1% (67.4%–96.8%) | 17.9% (3.2%–32.6%) |
| Not applicable<sup>†</sup> | 0 | - | - | - |
| Not stated<sup>†</sup> | 2 | - | - | - |
| **Highest Education Attainment** |  |  |  |  |
| University degree or higher | 35 | 10.3% (0.0%–23.6%) | 80.7% (65.6%–95.8%) | 9.1% (0.9%–17.3%) |
| Certificate/Diploma (non-degree) | 117 | 8.1% (3.1%–13.0%) | 81.8% (72.9%–90.6%) | 10.2% (2.8%–17.5%) |
| Currently studying (no qualification) | 17 | 27.3% (1.3%–53.2%) | 33.5% (7.3%–59.8%) | 39.2% (8.0%–70.4%) |
| Year 12 | 14 | 0.9% (0.0%–2.6%) | 96.4% (90.5%–100.0%) | 2.7% (0.0%–8.3%) |
| Year 10 or 11 | 21 | 0.0% (0.0%–0.0%) | 79.6% (58.0%–100.0%) | 20.4% (0.0%–42.0%) |
| Year 9 or below | 5 | 0.0% (0.0%–0.0%) | 93.8% (79.7%–100.0%) | 6.2% (0.0%–20.3%) |
| Don't know / rather not say | 1 | - | - | - |
| Not applicable<sup>†</sup> | 0 | - | - | - |
| Not stated<sup>†</sup> | 0 | - | - | - |
| Missing/Unknown<sup>†</sup> | 0 | - | - | - |
| **Service coverage** |  |  |  |  |
| Private dental insurance only | 71 | 11.3% (1.5%–21.1%) | 75.5% (60.7%–90.3%) | 13.2% (1.7%–24.8%) |
| Both private insurance and public eligibility | 15 | 17.4% (0.0%–38.9%) | 82.6% (61.1%–100.0%) | 0.0% (0.0%–0.0%) |
| Eligible for public dental care only | 62 | 5.8% (0.5%–11.2%) | 71.4% (57.0%–85.8%) | 22.8% (8.9%–36.7%) |
| No insurance / not eligible | 57 | 10.4% (0.0%–21.0%) | 83.6% (71.1%–96.1%) | 6.1% (0.0%–13.5%) |
| Missing | 5 | 0.0% (0.0%–0.0%) | 72.8% (31.2%–100%) | 27.2% (0.0%–68.8%) |
| **Residential Location** |  |  |  |  |
| Major cities | 117 | 9.2% (2.8%–15.7%) | 82.0% (72.9%–91.0%) | 8.8% (2.1%–15.5%) |
| Regional | 84 | 11.0% (3.1%–18.9%) | 67.7% (54.0%–81.4%) | 21.3% (8.3%–34.2%) |
| Remote/Very remote | 9 | 0.0% (0.0%–0.0%) | 38.3% (0.0%–81.0%) | 61.7% (19.0%–100%) |

**Notes:** Pathways reflect the highest-intensity service accessed: GP/Other (primary care), ED (non-admitted emergency department), and hospital admission. Sample size (n) reflects the unweighted number of respondents. Percentages represent population-weighted estimates with 95% confidence intervals derived from Taylor-linearised standard errors, accounting for the complex survey design. Results are stratified by demographic, socioeconomic, and access-to-care characteristics among respondents who used any medical service for a dental problem ( $n = 210$ ).
<sup>†</sup> Cells with fewer than 5 unweighted observations suppressed.

### Treatment Pathways within Non-dental Settings

Treatment varied markedly according to the reason for presentation (Table 3). Script prescribing was most common for toothache/wisdom-tooth problems (62.8%) and oral injuries (59.4%). Antibiotics dominated prescriptions for oral injuries (88.0%), consistent with trauma-related management. In contrast, only one in four adults presenting with other acute dental problems received any prescription (25.9%).

**Table 3.** Treatment Pathways Among Patients Seeking Medical Care for Dental Problems, by Reason for Presentation.

| Reason | Toothache/Wisdom tooth<br>(95% CI) | Oral Injury<br>(95% CI) | Other acute problem<br>(95% CI) |
| --- | --- | --- | --- |
| n (unweighted) | 96 | 22 | 65 |
| Script prescribed | 62.8% (48.1–77.5) | 59.4% (31.8–86.9) | 25.9% (7.2–44.5) |
| *Pain relief* | 31.2% (10.7–51.7) | 12.0% (0.0–40.5) | - |
| *Antibiotics* | 44.7% (23.9–65.4) | 88.0% (59.5–100.0) | 39.7% (0.9–78.6) |
| *Both* | 24.1% (5.7–42.5) | - | 18.2% (0.0–49.0) |
| *Don't know* | - | - | 42.0% (0.0–84.0) |
| No script | 37.2% (22.5–51.9) | 40.6% (13.1–68.2) | 74.1% (55.5–92.8) |
| Referred to dentist | 11.5% (0.0–33.6) | 19.9% (0.0–71.3) | 19.0% (0.0–43.6) |
| Received dental treatment | 64.3% (28.0–100.0) | 80.1% (28.7–100.0) | 1.4% (0.0–4.9) |
| Other medical | 7.8% (0.0–25.8) | - | - |
| Other | 8.0% (0.0–22.5) | - | 51.2% (17.9–84.6) |
| No treatment | 8.4% (0.0–23.9) | - | 28.4% (0.0–61.2) |

**Note:** Estimates are survey-weighted percentages with 95% confidence intervals, accounting for the complex survey design using Taylor-linearised variance estimation. Medication subcategories (pain relief, antibiotics, both, don't know) refer only to those who received a script. Treatment categories reflect self-reported care received during the medical visit and may include general medical procedures (e.g., wound care, irrigation, analgesic administration). Small unweighted subgroup sizes resulted in wide confidence intervals, and estimates should be interpreted with caution.

Non-pharmacological treatment also differed. Dental-type interventions were frequently reported for toothache/wisdom-tooth problems (64.3%) and oral injuries (80.1%) but were rarely provided for other acute problems (1.4%). Management of “other acute problems” was heterogeneous: 51.2% received general advice or non-specific treatment, 28.4% received no treatment, and only 1.4% received a dental intervention. Referral to a dentist was uncommon across all presentation types (11.5–19.9%).

### DISCUSSION

The National Dental Telephone Interview Survey (NDTIS) 2021 offers a detailed account of how dental problems are managed within Australian medical services and highlights the extent to which these pathways differ according to the nature of the presenting problem. Based on the survey’s weighted prevalence estimate (4.4%) and applying this proportion to the current Australian adult population (approximately 20 million adults aged 18 years and over) (ABS, 2023), this would equate to roughly 880,000 to 900,000 adults each year seeking medical care, rather than dental care, for a dental problem each year. Overall, our findings reinforce what clinicians have observed anecdotally for many years: people seek medical care for dental issues when access to a dentist is problematic, and the care they receive is highly variable, often resource-intensive, and not always aligned with best practice (AIHW, 2023b; Duckett et al., 2019). From a public health and medical education perspective, the picture that emerges is one of a system absorbing dental demand by default rather than by design.

Clear differences in treatment patterns were evident across the three presentation groups. Toothache and wisdom-tooth problems were associated with the highest level of antibiotic prescribing, with nearly two-thirds of patients receiving a script and almost half of these scripts being antibiotics. Oral injury presentations showed a similarly high level of prescribing, again dominated by antibiotics. These patterns are consistent with previous work shows that general practitioners frequently prescribe antibiotics for dental presentations (Cope et al., 2016), reflecting a wider reliance on antibiotics in the absence of definitive dental treatment, a pattern also documented within dental practice itself (Palmer et al., 2000). The finding that a substantial proportion of participants were unable to recall the type of medication they received (particularly in the “other acute problem” category) further suggests challenges in diagnostic clarity and communication during these medical encounters.

Non-pharmacological management also differed substantially by presentation type. Oral injury was overwhelmingly managed through procedural care, with approximately four in five participants reporting some form of “dental treatment.” In the Australian context, this likely reflects the acute management required for traumatic dental injuries, including wound care and stabilisation (Lam, 2016), which may account for the high proportion of procedural care. Toothache/wisdom tooth problems were also frequently managed procedurally, suggesting that clinicians often attempt some form of hands-on intervention when a patient presents with severe pain but cannot access a dentist. By contrast, other acute dental problems, an intentionally heterogeneous group, were managed through a wide mix of approaches, including advice only, symptomatic treatment, and, for some, no intervention at all. This pattern aligns with prior research showing high variability in how non-dental clinicians interpret and manage dental conditions (Akinlotan & Ferdinand, 2020).

The study’s findings provide a view of medical services as a “catch-all” provider for dental problems. In the absence of affordable, timely dental care, people are relying on GPs, urgent care centres, and emergency departments for issues that sit outside the core skill set of medical clinicians (Duckett et al., 2019). This pattern is not surprising given that dental care in Australia remains largely outside Medicare, and out-of-pocket costs are substantial (AIHW, 2023b). Individuals with limited financial resources often have little choice but to seek care from the parts of the health system that remain accessible to them; namely, bulk-billing medical clinics and public hospitals (AIHW, 2023b; Duckett et al., 2019). The high proportion of participants who present to emergency departments, and the small proportion who are subsequently referred to a dentist, speak to the fragmented pathways that people navigate when dental care is unavailable or unaffordable (Akinlotan & Ferdinand, 2020).

Our findings also raise important questions about medical education. Dental conditions receive very limited attention in GP training and even less in hospital rotations, with Australian medical graduates receiving little, if any, foundational knowledge of oral health and minimal exposure to dental diseases, trauma, or oral–systemic conditions (Abbott et al., 2018). As a result, antibiotic prescribing may function as a default strategy, particularly when clinicians feel they must provide “something” to the patient. The variability observed in advice-based or “other” treatment also suggests that clinicians may rely on general principles of pain management rather than evidence-based dental protocols. Collectively, these issues point to opportunities for clearer national guidance, targeted continuing education, and more structured referral pathways between medical and dental services.

The study has several strengths. It uses a large, nationally representative dataset and applies survey weighting to produce generalisable estimates. It also provides a level of treatment granularity that has not been described previously in Australian dental-medical utilisation research. However, findings should be interpreted with several limitations in mind. All data are self-reported, and participants may have had difficulty recalling the exact treatment or medication received, contributing to the sizeable “don’t know” and “other” categories. Some may also have interpreted general medical procedures, such as wound care, irrigation, or analgesic administration, as “dental treatment,” potentially overstating the extent of dental-type care provided in medical settings. The dataset does not distinguish between specific clinical contexts. A single episode may involve both a GP and an emergency department, yet these are recorded together, meaning some pathways, particularly for oral injury, likely reflect a combination of ED trauma management and hospital dental involvement. Finally, small subgroups resulted in wide confidence intervals, and these estimates should be seen as indicative rather than precise. Nonetheless, the overall patterns remain consistent and align with broader evidence on dental care access in Australia.

## CONCLUSION

At a system level, the survey reflects deeper structural and policy issues. That so many individuals resort to emergency departments for dental care signals gaps in preventive and primary dental service availability. It also places unnecessary pressure on ED resources and leads to repeated cycles of temporary relief followed by symptom recurrence, since definitive treatment is rarely provided in medical settings. Strengthening dental service accessibility, whether through public dental expansion, integrated medical–dental clinics, or targeted Medicare-funded urgent dental care, would likely reduce the burden on medical services while ensuring that patients receive appropriate care earlier in the course of their condition. Policies that facilitate direct pathways from EDs or GPs to dental clinics, such as guaranteed urgent dental appointments or co-located services, may also reduce repeated medical presentations.

## ACKNOWLEDGMENTS

The National Dental Telephone Interview Survey (NDTIS) 2021 was funded by the Australian Government Department of Health and Aged Care (DoH). The authors would like to acknowledge all those who participated in the Survey.

## Declarations

### Ethics approval

The NDTIS 2021 was approved by the Human Research Ethics Committee at the University of Adelaide (approval number H-2020–153).

### Consent to participate

Participants provided their informed consent prior to survey completion. All datasets were de-identified to ensure anonymity.

### Consent for publication

Not applicable.

### Availability of data and material

The datasets generated and/or analysed during the current study are not publicly available. We do not have permission from the University of Adelaide Human Research Ethics Committee to publicly release the dataset in either identifiable or de-identified form.

### Code availability

The R syntax is available from the corresponding author upon reasonable request.

### Authors’ contributions

All authors contributed to the study conception and design of this manuscript. Material preparation, data collection and analysis were performed by Sergio Chrisopoulos, Liana Luzzi, and Kym McCormick. The first draft of the manuscript was written by Liana Luzzi and all authors commented on previous versions of the manuscript. All authors read and approved the final manuscript.

## REFERENCES

Abbott, B., Zybutz, C., Scott, K. M., Eberhard, J., & Widmer, R. (2018). A review of the hours dedicated to oral health education in medical programmes across Australia. Internal Medicine Journal, 48, 1035–1040. 10.1111/imj.14021

Akinlotan, M. A., & Ferdinand, A. O. (2020). Emergency department visits for nontraumatic dental conditions: a systematic literature review. J Public Health Dent, 80(4), 313–326. 10.1111/jphd.12386

Australian Bureau of Statistics. (2023). National, state and territory population. https://www.abs.gov.au/statistics/people/population/national-state-and-territory-population/dec-2023.

Australian Institute of Health and Welfare (AIHW). (2023a). AIHW Hospital Morbidity Database: 2016–17 to 2020–21.

Australian Institute of Health and Welfare (AIHW). (2023b). Health expenditure Australia 2022-23. https://www.aihw.gov.au/reports/health-welfare-expenditure/health-expenditure-australia

Brennan, D. S., Luzzi, L., Chrisopoulos, S., & Haag, D. G. (2020). Oral health impacts among Australian adults in the National Study of Adult Oral Health (NSAOH) 2017–18. Australian Dental Journal, 65, S59–S66. 10.1111/adj.12766

Cope, A. L., Chestnutt, I. G., Wood, F., & Francis, N. A. (2016). Dental consultations in UK general practice and antibiotic prescribing rates: a retrospective cohort study. British Journal of General Practice. 10.3399/bjgp16X684757

Davis, E. E., Deinard, A. S., & Maïga, E. W. (2010). Doctor, my tooth hurts: the costs of incomplete dental care in the emergency room. Journal of public health dentistry, 70, 205–210. 10.1111/j.1752-7325.2010.00166.x

Duckett, S., Cowgill, M., & Swerissen, H. (2019). Filling the gap: a universal dental care scheme for Australia. Grattan Institute. https://grattan.edu.au/wp-content/uploads/2019/03/915-Filling-the-gap-A-universal-dental-scheme-for-Australia.pdf

Hopcraft, M. (2024). Commentary on the senate select committee into the provision of and access to dental services in Australia: an opportunity for reform. Australian Dental Journal, 69(3), 162–174. 10.1111/adj.13012

Lam, R. (2016). Epidemiology and outcomes of traumatic dental injuries: a review of the literature. Australian Dental Journal, 61, 4–20. 10.1111/adj.12395

Lee, H. H., Lewis, C. W., Saltzman, B., & Starks, H. (2012). Visiting the emergency department for dental problems: trends in utilization, 2001 to 2008. Am J Public Health, 102(11), e77–83. 10.2105/ajph.2012.300965

Lumley, T. (2004). Analysis of complex survey samples. Journal of statistical software, 9, 1–19. 10.18637/jss.v009.i08

National Advisory Council on Dental Health. (2012). Report of the National Advisory Council on Dental Health Dept. of Health and Ageing. https://catalogue.nla.gov.au/catalog/5978887

Palmer, N. O., Martin, M. V., Pealing, R., & Ireland, R. S. (2000). An analysis of antibiotic prescriptions from general dental practitioners in England. Journal of antimicrobial chemotherapy, 46(6), 1033–1035. 10.1093/jac/46.6.1033

Rubin, D. B. (1996). Multiple imputation after 18+ years. Journal of the American statistical Association, 91(434), 473–489. 10.1080/01621459.1996.10476908

Sterne, J. A. C., White, I. R., Carlin, J. B., Spratt, M., Royston, P., Kenward, M. G., Wood, A. M., & Carpenter, J. R. (2009). Multiple imputation for missing data in epidemiological and clinical research: potential and pitfalls. BMJ, 338, b2393. 10.1136/bmj.b2393

Van Buuren, S., & Groothuis-Oudshoorn, K. (2011). mice: Multivariate imputation by chained equations in R. Journal of statistical software, 45, 1–67. 10.18637/jss.v045.i03

Zou, G. (2004). A modified poisson regression approach to prospective studies with binary data. American journal of epidemiology, 159(7), 702–706. 10.1093/aje/kwh090

